# Pembrolizumab in advanced acral lentiginous melanoma: final results of a single-centre, open-label, phase II trial in an East Asian population

**DOI:** 10.64898/2026.07.22.26358641

**Authors:** Herbert H. Loong, Winnie Yeo, Candy Yuen, Frankie Mo, Tung-Ching Chan, Kirsty W. C. Lee, Alvita C. Y. Chan, Ashley C. Y. Wong, Kenneth C. W. Wong, Daisy C. M. Lam, Joanna Tong, Carlos K. H. Wong

**Author notes:** **Correspondence:** Dr. Herbert H. Loong, Department of Clinical Oncology, The Chinese University of Hong Kong, Prince of Wales Hospital, Shatin, Hong Kong SAR. **Trial registration:** ClinicalTrials.gov NCT02875132 (institutional code MEL001).

## Abstract

**Background:** Acral lentiginous melanoma (ALM) is the predominant melanoma subtype in East Asian populations, accounting for roughly 50–58% of cases, compared with 2–3% in populations of European ancestry. ALM is genomically and biologically distinct from sun-exposed cutaneous melanoma, and East Asian and acral patients were markedly under-represented in the pivotal anti–PD-1 registration trials. At the time this study was designed, no prospective trial had evaluated a checkpoint inhibitor specifically in ALM. We conducted a phase II trial to estimate the activity of pembrolizumab in this population.

**Methods:** In this single-centre, single-arm, open-label phase II trial, adults with metastatic or locoregionally advanced inoperable ALM who were naïve to anti–PD-1/PD-L1 therapy received pembrolizumab 200 mg intravenously every 3 weeks until progression, unacceptable toxicity, or withdrawal. The primary endpoint was objective response rate (ORR) by RECIST 1.1. Secondary endpoints included duration of response (DoR), clinical benefit rate (CBR), progression-free survival (PFS), overall survival (OS), and safety (CTCAE v4.0). A Simon minimax two-stage design (P0=0.10, P1=0.30, α=0.05, power=80%) planned enrolment of up to 28 patients.

**Results:** Between February 2017 and June 2019, 9 patients were enrolled before recruitment was halted for slow accrual, the interval availability of reimbursed pembrolizumab, and a low observed response signal. Median age was 72 years (range 48–78); 6 (67%) were male; all had ECOG performance status 0 and metastatic disease; 7 (78%) had received prior therapy. One patient achieved a partial response (ORR 11.1%, 95% CI 0.0–31.6%), with a DoR of 19 months; 3 had stable disease and 4 progressed. CBR (response or stable disease ≥12 weeks) was 44.4% (95% CI 12.0–76.9). At a median follow-up of 7.6 months, median PFS was 3.4 months (95% CI 1.4–21.3) and median OS was 7.6 months (95% CI 2.0– 34.3). Two grade 3 adverse events occurred, both assessed as unrelated to study drug; no treatment-related grade ≥3 events were recorded. In an exploratory analysis, an LDH-to–upper-limit-of-normal ratio >1.5 was associated with worse OS (median 4.3 vs 26.5 months; HR 4.58, 95% CI 0.82–25.7; log-rank p=0.06).

**Conclusions:** Recruitment was constrained by disease rarity and a shifting reimbursement landscape, and the trial closed before completing stage I. Within these limitations, single-agent pembrolizumab showed only modest activity in advanced ALM, consistent with the limited efficacy subsequently reported in larger contemporary acral melanoma cohorts. The exploratory association between elevated LDH ratio and poorer survival warrants prospective evaluation.

**What is already known / what this study adds:**

- ALM is the commonest melanoma subtype in East Asia but was almost absent from the trials that established PD-1 blockade; no prospective checkpoint-inhibitor trial in pure ALM existed when this study was designed.
- In this prospective phase II trial, single-agent pembrolizumab produced an ORR of 11.1% with modest survival, consistent with limited activity later reported in larger acral cohorts.
- An exploratory signal linking elevated LDH ratio to poorer survival supports its prognostic relevance and the need for collaborative, biomarker-driven trials dedicated to ALM.

## 1. Introduction

Cutaneous melanoma is broadly divided into subtypes that differ in anatomical distribution, ultraviolet aetiology, and driver mutation landscape. Acral lentiginous melanoma (ALM) arises on the palms, soles, and nail apparatus—skin that is not chronically sun-exposed—and is defined by its distinctive histopathology. Although ALM constitutes only 2–3% of melanoma in populations of European ancestry, it is the single most common subtype among East Asian patients, where it accounts for approximately 50– 58% of cases in reported series. This epidemiological inversion means that a disease considered rare in the West is, in this locality, the melanoma a medical oncologist encounters most often.

ALM is not simply cutaneous melanoma at an unusual site. Genomic studies have shown that acral melanomas carry a distinct pattern of genetic alterations, with a lower burden of ultraviolet-signature mutations, a lower frequency of BRAF V600 mutations, and more frequent structural aberrations and gene amplifications than sun-related cutaneous melanoma. These biological differences raise a legitimate question about whether efficacy data generated predominantly in patients with sun-exposed cutaneous melanoma can be extrapolated to ALM.

This concern is compounded by the composition of the pivotal anti–PD-1 trials. In KEYNOTE-001, which established the activity of pembrolizumab in advanced melanoma, Asian patients comprised only about 2% of the study population, and acral cases were correspondingly scarce. When this study was conceived, no prospective clinical trial had evaluated a checkpoint inhibitor specifically in patients with ALM, and the applicability of existing immunotherapy data to this population was essentially unknown. We therefore undertook a prospective phase II trial to estimate the objective response rate of pembrolizumab in biological-therapy–naïve patients with advanced ALM, together with survival and safety outcomes, in a predominantly East Asian population in Hong Kong.

## 2. Methods

### 2.1 Study design and oversight

This was a single-centre, single-arm, open-label, phase II clinical trial (institutional code MEL001; ClinicalTrials.gov NCT02875132) sponsored by the Comprehensive Cancer Trial Centre, Department of Clinical Oncology, The Chinese University of Hong Kong, and conducted at the Phase 1 Clinical Trials Centre of the same institution. The trial was activated in February 2017. The protocol and all amendments were approved by the institutional research ethics committee, and the trial was conducted in accordance with the Declaration of Helsinki and Good Clinical Practice guidelines. All patients provided written informed consent before any study procedure.

### 2.2 Patients

Eligible patients were adults (≥18 years) with a histologically confirmed diagnosis of metastatic or locoregionally advanced and inoperable acral lentiginous melanoma, measurable disease by RECIST 1.1, and an ECOG performance status of 0 or 1. Patients were required to provide tumour tissue from a newly obtained or archival biopsy for correlative studies. Key exclusion criteria were prior therapy with an anti– PD-1, anti–PD-L1, or anti–PD-L2 agent; systemic anticancer therapy or radiotherapy within 2 weeks of the first study dose, or unresolved toxicity from prior treatment; active autoimmune disease requiring systemic treatment within the preceding 2 years; a history of or active non-infectious pneumonitis; and known HIV, active hepatitis B, or active hepatitis C infection. Prior cytotoxic chemotherapy or molecularly targeted therapy was permitted.

### 2.3 Treatment

Patients received pembrolizumab 200 mg by intravenous infusion every 3 weeks. Treatment continued until documented disease progression, unacceptable toxicity, withdrawal of consent, or investigator decision; patients with radiographic progression who were judged to be deriving clinical benefit could continue treatment at the investigator’s discretion, with progression confirmed on repeat imaging at least 4 weeks later. Dose reductions were not permitted; dose delays and interruptions for toxicity were managed per protocol. Adverse events were graded according to the Common Terminology Criteria for Adverse Events (CTCAE) version 4.0.

### 2.4 Endpoints and assessments

The primary endpoint was objective response rate (ORR), defined as the proportion of patients achieving a complete or partial response by RECIST 1.1. Secondary endpoints were duration of response, clinical benefit rate (CBR), progression-free survival (PFS), overall survival (OS), response assessment by immune-related response criteria (irRC), and safety and tolerability. CBR was defined as complete response, partial response, or stable disease sustained for a pre-specified interval; results are reported for both a ≥6-month and a ≥12-week stable-disease threshold. Radiological tumour assessments were performed by computed tomography and evaluated per RECIST 1.1, with reassessment every two cycles during the first 24 months. Pre-specified exploratory objectives included baseline and on-treatment tumour PD-L1 expression and pharmacokinetic characterisation of pembrolizumab in an East Asian population; these correlative analyses were not completed owing to early trial closure. A separate exploratory analysis examined the association between the ratio of serum lactate dehydrogenase (LDH) to the upper limit of normal and survival, dichotomised at a ratio of 1.5.

### 2.5 Statistical analysis

A Simon minimax two-stage design was used to determine sample size. Pembrolizumab was considered inactive if the true ORR was ≤10% (P0=0.10) and worthy of further study if the ORR was ≥30% (P1=0.30), with a one-sided type I error of 0.05 and 80% power. Under this design, an initial cohort was to be accrued in stage I, with expansion to a planned total of up to 28 patients if a pre-specified activity threshold was met. Response and clinical benefit rates are presented with exact (Clopper–Pearson) 95% confidence intervals. PFS and OS were estimated by the Kaplan–Meier method. In the exploratory subgroup analysis, OS was compared by the log-rank test and hazard ratios were derived from a Cox proportional-hazards model. All patients who received at least one dose of pembrolizumab were included in the efficacy and safety analyses.

## 3. Results

### 3.1 Patients and recruitment

The first patient was enrolled on 17 February 2017 and the last on 27 June 2019. Over this 28-month period, 9 patients were enrolled—far short of the planned accrual. Recruitment was formally halted in June 2019 for three converging reasons: the rarity of the disease produced persistently slow accrual; pembrolizumab became available through reimbursement during the enrolment window, reducing the incentive for eligible patients to enrol in a trial; and only 1 of the first 9 patients had achieved an objective response, suggesting limited single-agent activity. The trial therefore closed before completing the planned stage I cohort, and the results below are descriptive.

Baseline characteristics are summarised in Table 1. Median age was 72 years (range 48–78); 6 patients (67%) were male. All 9 patients had an ECOG performance status of 0 and all had metastatic disease at study entry. Four patients (44%) had a single metastatic site and 5 (56%) had two or more sites. Seven patients (78%) had received prior treatment of some form—including prior surgery in 7 (78%), radiotherapy in 4 (44%), and cytotoxic chemotherapy in 2 (22%)—while 2 (22%) were treatment-naïve. Eight patients (89%) had acral lentiginous histology; one had a melanoma of the right great toe with nodal metastasis.

**Table 1.** Baseline demographic and clinical characteristics (N=9).

| Characteristic | Category | N (%) |
| --- | --- | --- |
| Age, years | Median (range) | 72 (48–78) |
|  | Mean (SD) | 68 (9.9) |
| Sex | Male | 6 (66.7) |
|  | Female | 3 (33.3) |
| ECOG performance status | 0 | 9 (100) |
| Histology | Acral lentiginous melanoma | 8 (88.9) |
|  | Melanoma, right great toe with nodal mets | 1 (11.1) |
| Disease status | Metastatic | 9 (100) |
| Number of metastatic sites | 1 | 4 (44.4) |
|  | 2 | 2 (22.2) |
|  | 3 | 1 (11.1) |
|  | 4 | 2 (22.2) |
| Any prior treatment | Yes | 7 (77.8) |
| Prior surgery | Yes | 7 (77.8) |
| Prior radiotherapy | Yes | 4 (44.4) |
| Prior chemotherapy | Yes | 2 (22.2) |
| Stage at initial diagnosis | I / II / III / IV | 1 / 6 / 1 / 1 |

SD, standard deviation; ECOG, Eastern Cooperative Oncology Group; mets, metastasis. Percentages may not sum to 100 owing to rounding.

### 3.2 Treatment exposure

Patients received a median of 6 cycles of pembrolizumab (range 2–36; mean 10.8, SD 10.9). One patient (PW003) had a protocol-specified dose delay and dose interruption at the investigator’s discretion; no dose reductions occurred. At data cut-off, all patients had discontinued treatment, and in every case the reason for discontinuation was disease progression.

### 3.3 Efficacy

All 9 patients received at least one dose of pembrolizumab and were evaluable for safety; efficacy is reported for the same population, of whom 8 had an evaluable radiological response (one patient was not evaluable). One patient achieved a partial response, for an ORR of 11.1% (95% CI 0.0–31.6%). Three patients had stable disease and 4 had progressive disease as their best response. Best percentage change in target lesions and best overall response for each patient are shown in Table 2. The single responder (PW007) had a deep response, with a 56% reduction in target-lesion burden and a duration of response of 588 days (approximately 19 months).

**Table 2.** Best overall response and best change in target lesion by patient.

| Patient | Best change in target lesion (%) | Best response | Cycles received |
| --- | --- | --- | --- |
| PW001 | +1.9 | PD | 6 |
| PW002 | +20.0 | SD | 6 |
| PW003 | –26.7 | SD | 32 |
| PW004 | Not evaluable | NE | 2 |
| PW005 | +94.7 | PD | 6 |
| PW006 | +31.8 | PD | 3 |
| PW007 | –56.2 | PR | 36 |
| PW008 | +28.6 | SD | 5 |
| PW009 | Not evaluable | PD | 2 |

PR, partial response; SD, stable disease; PD, progressive disease; NE, not evaluable. Response assessed per RECIST 1.1.

Clinical benefit rate depended on the stable-disease threshold applied. Using complete or partial response, or stable disease lasting at least 6 months, CBR was 22.2% (95% CI 0.0–49.4). Using a less stringent threshold of stable disease lasting at least 12 weeks, CBR was 44.4% (95% CI 12.0–76.9).

### 3.4 Survival

At a median follow-up of 7.6 months (range 2.0–34.3), all 9 patients had experienced disease progression and 1 patient remained alive. Median progression-free survival was 3.4 months (95% CI 1.4–21.3) and median overall survival was 7.6 months (95% CI 2.0–34.3) (Figures 2 and 3).

**Figure 1.**
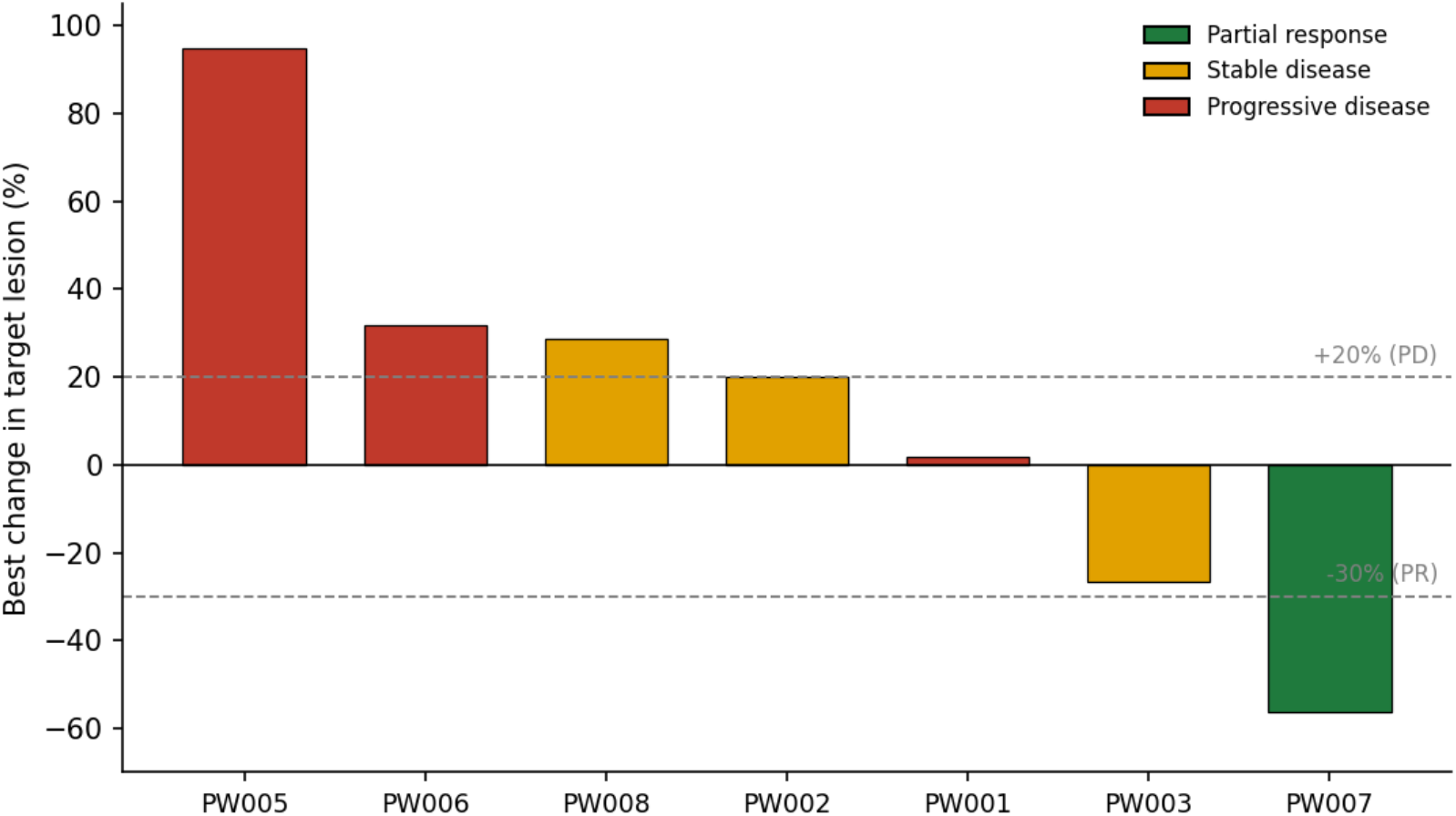
Waterfall plot of best percentage change in target-lesion burden for the 7 patients with an evaluable response. Bars are coloured by best overall response (RECIST 1.1). Dashed lines denote the +20% progression and −30% response thresholds. Two patients (PW004, PW009) were not evaluable and are not shown.

**Figure 2.**
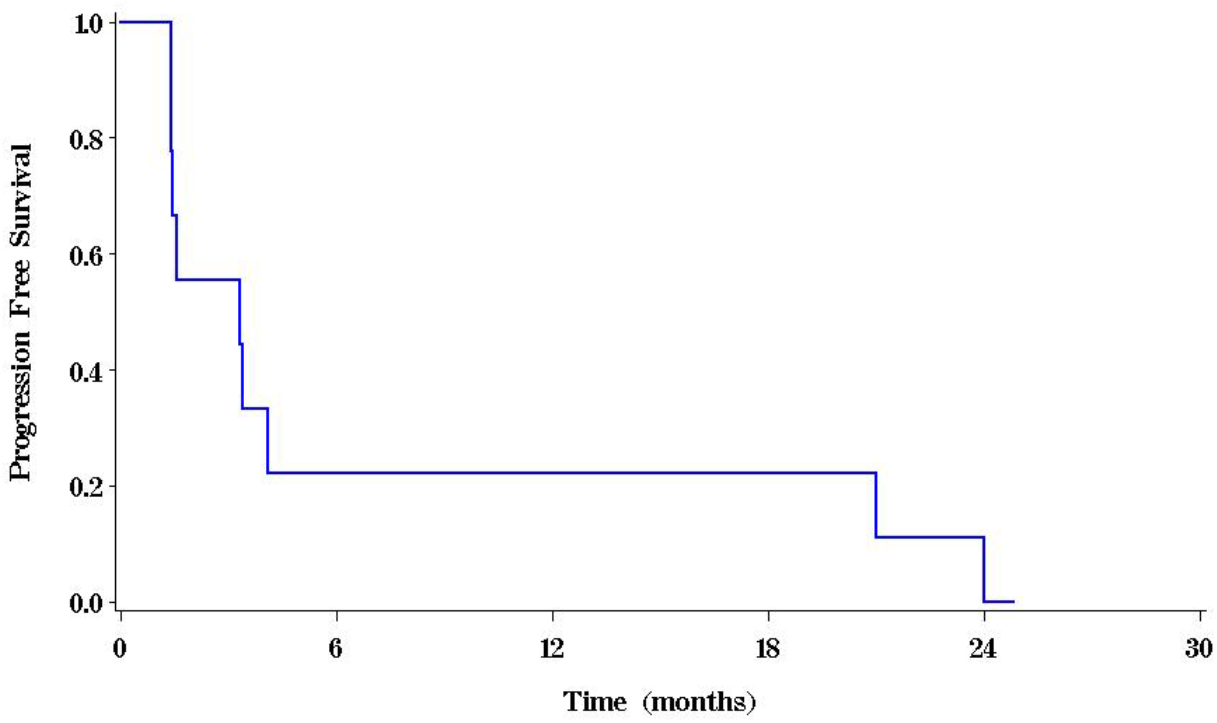
Kaplan–Meier estimate of progression-free survival for the full analysis population (N=9). Median progression-free survival was 3.4 months (95% CI 1.4–21.3).

**Figure 3.**
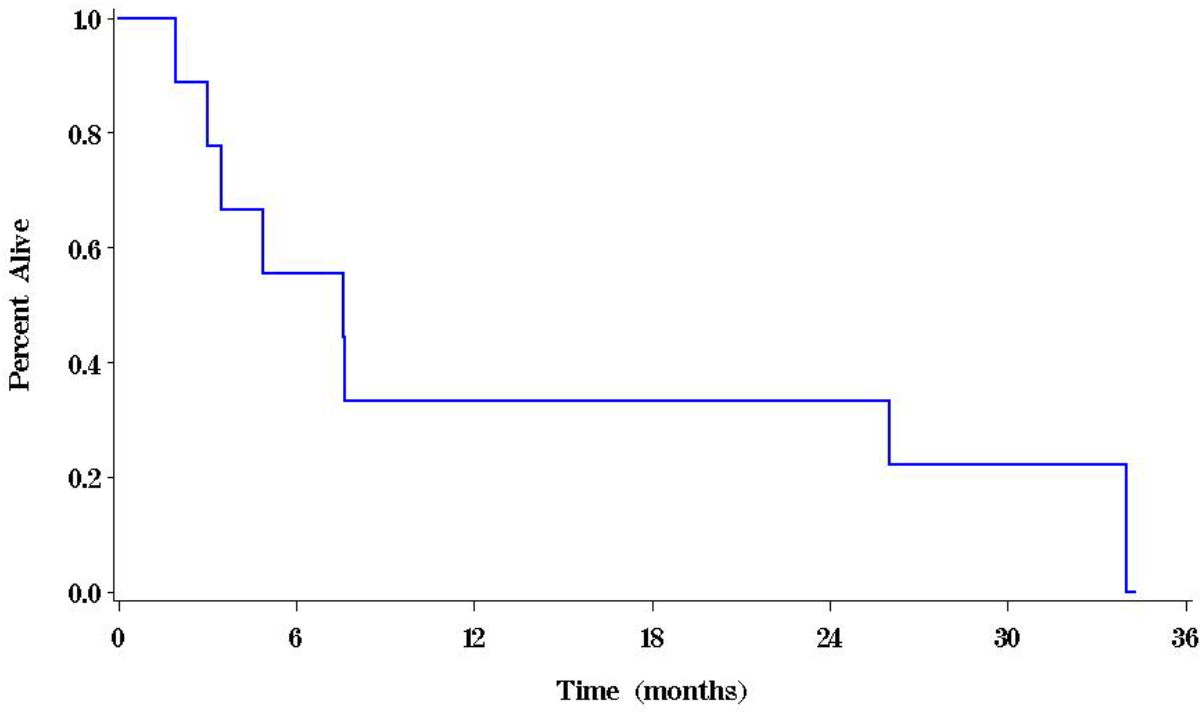
Kaplan–Meier estimate of overall survival for the full analysis population (N=9). Median overall survival was 7.6 months (95% CI 2.0–34.3).

### 3.5 Safety

Pembrolizumab was well tolerated. Reported adverse events were predominantly grade 1 and included cough, malaise, nausea, epigastric pain, and skin rash (Table 3). Two grade 3 adverse events were recorded—dizziness in one patient (PW008) and pleural effusion in another (PW009)—both of which were assessed by the investigator as unrelated to study drug and both of which resolved. No treatment-related grade 3 or higher adverse events and no treatment-related deaths occurred. The grade 3 pleural effusion in PW009 led to regimen discontinuation but was considered unrelated to pembrolizumab and occurred in the setting of progressive disease.

**Table 3.** Treatment-emergent adverse events by maximum grade (N=9).

| Adverse event | Grade 1 | Grade 2 | Grade 3 | Total |
| --- | --- | --- | --- | --- |
| Dizziness | 0 | 0 | 1 | 1 |
| Pleural effusion | 0 | 0 | 1 | 1 |
| Cough | 1 | 0 | 0 | 1 |
| Malaise | 1 | 0 | 0 | 1 |
| Nausea | 1 | 0 | 0 | 1 |
| Epigastric pain | 1 | 0 | 0 | 1 |
| Skin rash | 1 | 0 | 0 | 1 |

*Graded per CTCAE v4.0. Both grade 3 events (dizziness, pleural effusion) were assessed as unrelated to pembrolizumab; no treatment-related grade $\geq 3$ events occurred.*

### 3.6 Exploratory analysis: LDH ratio and survival

An exploratory analysis dichotomised patients by the ratio of serum LDH to the upper limit of normal at a cut-off of 1.5 (Table 4). The 4 patients with an LDH ratio >1.5 had a markedly shorter median overall survival than the 5 patients with a ratio ≤1.5 (4.3 vs 26.5 months). The corresponding hazard ratio was 4.58 (95% CI 0.82–25.7), with a log-rank p-value of 0.06. Although this difference did not reach statistical significance—unsurprising given the very small sample—the magnitude and direction of the effect are consistent with the well-established prognostic role of LDH in melanoma and merit prospective confirmation.

**Table 4.** Overall survival by LDH-to–upper-limit-of-normal ratio.

| LDH ratio | N | Median OS, months (95% CI) | HR (95% CI) | p-value |
| --- | --- | --- | --- | --- |
| $\leq 1.5$ | 5 | 26.5 (4.9–34.3) | 1 (ref) | — |
| $> 1.5$ | 4 | 4.3 (2.0–11.8) | 4.58 (0.82–25.7) | 0.06* |

OS, overall survival; HR, hazard ratio; CI, confidence interval; LDH, lactate dehydrogenase. \*Log-rank p-value; the Cox model p-value was 0.08. Analysis is exploratory and hypothesis-generating.

**Figure 4.**
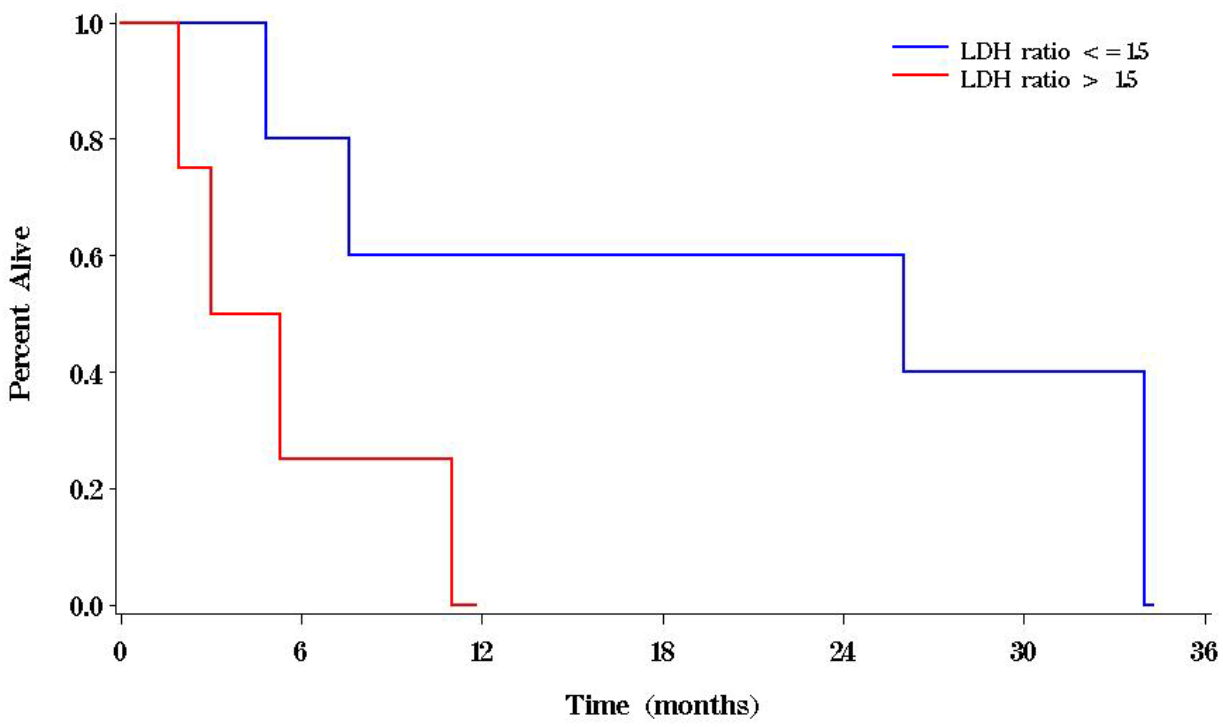
Kaplan–Meier estimates of overall survival by LDH ratio. Patients with an LDH-to–upper-limit-of-normal ratio ≤1.5 (blue; n=5) had longer overall survival than those with a ratio >1.5 (red; n=4): median 26.5 vs 4.3 months (HR 4.58, 95% CI 0.82– 25.7; log-rank p=0.06).

## 4. Discussion

This prospective phase II trial set out to estimate the activity of single-agent pembrolizumab in advanced acral lentiginous melanoma, a subtype that dominates the melanoma caseload in East Asia yet was almost absent from the trials that defined checkpoint-inhibitor practice. The trial did not meet its accrual goal: only 9 of a planned 28 patients were enrolled before recruitment was stopped. Any conclusions must therefore be interpreted with caution. Within these constraints, the observed objective response rate of 11.1% falls at or below the boundary the design defined as consistent with an inactive agent (P0=10%), and well below the 30% target that would have justified expansion. Single-agent pembrolizumab, in this small cohort, showed only modest activity.

These results should be read alongside the larger body of evidence that has accumulated since the trial was designed—much of it reinforcing the same message. In the Chinese KEYNOTE-151 study, second-line pembrolizumab produced an objective response rate of about 16% in the acral subtype. A multi-institutional retrospective series by Shoushtari and colleagues reported a higher acral response rate of roughly 32% with anti–PD-1 agents, but subsequent and larger cohorts have generally been less encouraging: a multicentre study of 193 Japanese acral melanoma patients found limited efficacy of anti– PD-1 monotherapy, and pooled retrospective data have converged on response rates in the mid-teens with median overall survival broadly in the range of 18–26 months. Registry analyses of first-line checkpoint inhibition in metastatic ALM have likewise reported outcomes inferior to those seen in non-acral cutaneous melanoma. Our finding of limited single-agent activity is therefore concordant with the contemporary consensus that ALM responds less well to PD-1 blockade than sun-exposed cutaneous melanoma.

Several biological features may underlie this relative resistance. Acral melanomas carry a lower tumour mutational burden and fewer ultraviolet-signature mutations than sun-exposed cutaneous melanoma, yielding fewer neoantigens for immune recognition; they more often harbour structural copy-number changes and amplifications; and features of the acral tumour microenvironment may be less permissive to checkpoint-inhibitor activity. Together these differences provide a plausible mechanistic basis for the lower response rates observed across acral cohorts, and they argue that ALM should be studied—and treated—as a biologically distinct entity rather than as a variant of common cutaneous melanoma.

Despite the limited overall activity, it is worth noting that the single responder in our cohort had a deep and durable response lasting approximately 19 months, and that one patient remained alive at last follow-up. This pattern—a minority of patients deriving substantial, durable benefit against a background of limited average activity—is characteristic of immunotherapy and underscores the need for predictive biomarkers to identify the patients most likely to benefit. Our exploratory observation that an elevated LDH ratio (>1.5 × upper limit of normal) was associated with substantially shorter survival is consistent with the established prognostic role of LDH in melanoma and, although not statistically significant in this small sample, is a hypothesis worth testing in adequately powered acral cohorts. The planned PD-L1 and pharmacokinetic correlatives, which could have refined patient selection and dosing in this population, were unfortunately not realised because of early closure.

The principal limitation of this study is its small size and premature closure. Beyond the intrinsic rarity of ALM in an absolute sense, accrual was further undermined by a change in the treatment landscape during the enrolment period: as pembrolizumab became reimbursable, patients could access the drug without enrolling in a trial, removing much of the practical incentive to participate. This experience illustrates a broader challenge in conducting rigorous prospective trials in rare melanoma subtypes, and argues strongly for multicentre and international collaboration to reach adequate sample sizes. Additional limitations include the single-arm design, the single-centre setting, and the heterogeneity of prior treatment. The strengths of the study are its prospective design, its focus on a pure ALM population defined histologically, and its complete follow-up for the primary and survival endpoints.

## 5. Conclusion

In this prospective but prematurely closed phase II trial, single-agent pembrolizumab demonstrated only modest activity in patients with advanced acral lentiginous melanoma, with an objective response rate of 11.1% and a median overall survival of 7.6 months. Although the small sample size precludes a definitive efficacy estimate, the findings are consistent with the limited activity of PD-1 blockade subsequently reported in larger contemporary acral melanoma cohorts, and they reinforce the view that ALM is a distinct entity requiring dedicated therapeutic strategies. The exploratory association between an elevated LDH ratio and poorer survival warrants prospective evaluation. Adequately powered, collaborative studies—ideally testing combination or novel immunotherapeutic approaches—are needed to improve outcomes in this under-served population.

## Data Availability

All data produced in the present study are available upon reasonable request to the authors

## 6. Declarations

### Ethics approval and consent to participate

The trial was approved by the institutional research ethics committee of The Chinese University of Hong Kong and was conducted in accordance with the Declaration of Helsinki and Good Clinical Practice. All patients provided written informed consent.

### Consent for publication

Not applicable; no individually identifiable patient data are presented.

### Data availability

The de-identified data supporting the findings of this study may be made available from the corresponding author on reasonable request, subject to institutional and ethics-committee approval.

### Author contributions

HHL conceived and designed the study, was principal investigator, enrolled and treated patients, interpreted the data, and drafted the manuscript. WY, CY, T-CC, KWCL, ACYC, ACYW, KCWW, DCML enrolled patients into the study. FM, CKHW provided statistical support. JT provided pathology support. All authors approved this manuscript.

## Acknowledgements

The authors thank the patients and their families, and the staff of the Comprehensive Cancer Trial Centre and Phase 1 Clinical Trial Centre, The Chinese University of Hong Kong.

*Note: this reference list is provided as a working bibliography; citations (particularly the ADOREG registry report) should be verified and reformatted to the target repository/journal style before deposition.*

